# Hypothalamus volumes in adolescent Myalgic Encephalomyelitis/Chronic Fatigue Syndrome: Impact of self-reported fatigue and illness duration

**DOI:** 10.1101/2023.05.16.23290031

**Authors:** Hollie Byrne, Elisha K Josev, Sarah J Knight, Adam Scheinberg, Katherine Rowe, Lionel Lubitz, Marc L Seal

## Abstract

Adolescent Myalgic Encephalomyelitis/Chronic Fatigue Syndrome (ME/CFS) is a complex illness of unknown aetiology. Emerging theories suggest ME/CFS may reflect a progressive, aberrant state of homeostasis caused by disturbances within the hypothalamus, yet few studies have investigated this using magnetic resonance imaging in adolescents with ME/CFS. We conducted a volumetric analysis to investigate whether whole and regional hypothalamus volumes in adolescents with ME/CFS differed compared to healthy controls, and whether these volumes were associated with fatigue severity and illness duration. 48 adolescents (25 ME/CFS, 23 controls) were recruited. Lateralised whole and regional hypothalamus volumes, including the anterior–superior, superior tubular, posterior, anterior-inferior and inferior tubular subregions, were calculated from T1-weighted images. When controlling for age, sex and intracranial volume, Bayesian linear regression revealed no evidence for differences in hypothalamus volumes between groups. However, in the ME/CFS group, a negative linear relationship between right anterior-superior volumes and fatigue severity was identified, which was absent in controls. In addition, Bayesian ordinal regression revealed a likely-positive association between illness duration and right superior tubular volumes in the ME/CFS group. While these findings suggest overall comparability in regional and whole hypothalamus volumes between adolescents with ME/CFS and controls, preliminary evidence was identified to suggest greater fatigue and longer illness duration were associated with greater right anterior-superior and superior-tubular volumes, respectively. These regions contain the anterior and superior divisions of the paraventricular nucleus, involved in the neuroendocrine response to stress, suggesting involvement in ME/CFS pathophysiology. However, replication in a larger, longitudinal cohort is required.

## Introduction

Adolescent Myalgic Encephalomyelitis, also referred to as Chronic Fatigue Syndrome (ME/CFS), is a debilitating illness of unknown aetiology. Adolescents with ME/CFS present with severe, debilitating fatigue that persists for a minimum of three months, worsening of symptoms following periods of exertion, and a range of additional symptoms reflecting cognitive, sleep, immune, autonomic and neuroendocrine disturbances (Jason et al., 2006). While the aetiological mechanisms are unknown, recent theories suggest ME/CFS may reflect a progressive, aberrant state of homeostasis (Hatziagelaki et al., 2018; Mackay and Tate, 2018; Marks, 2023; Nacul et al., 2020; Tate et al., 2022), either as a cause or consequence of central nervous system (CNS) disturbances. These disturbances have typically been investigated via magnetic resonance imaging (MRI), but studies have primarily focused on adults, who may exhibit differences with adolescents in terms of symptomatology (Collin et al., 2015), prognosis (Devendorf et al., 2019), and responses to treatment (Crawley, 2017). Given the significant social and neurodevelopmental changes that occur in adolescence, it is unclear whether neuroimaging findings in adults can be translated to adolescents with ME/CFS (Josev et al., 2020), contributing to the limited understanding of this complex illness during this age period.

It has been suggested the aberrant state of homeostasis in ME/CFS may be linked to neuroinflammation or disturbances within the hypothalamus (Hatziagelaki et al., 2018; Mackay and Tate, 2018; Tate et al., 2022). The hypothalamus is subdivided into approximately a dozen nuclei (depending on segmentation criteria), each with specialised functions involved in homeostasis of neuroendocrine, behavioural, and autonomic processes (Shapiro et al., 2022). Such functions include nuclei within the anterior hypothalamus that facilitate thermoregulation and sleep, posterior nuclei that promote arousal and wakefulness, and tuberal nuclei that regulate the stress response, energy balance and pain (Chen et al., 2019). Functional asymmetries within the hypothalamus have also been documented (Kiss et al., 2020), with the left hypothalamus more dominant in regulating circadian rhythm, thyroid activity, thermoregulation and immune responses, and the right more dominant in modulating cardiovascular responses to stress (Xavier et al., 2009). Because of its many functions, the hypothalamus is implicated in a large number of disorders, including sleep (Desseilles et al., 2008), eating (Thomas et al., 2019) and mood (Schindler et al., 2019) disorders, as well as progressive illnesses such as dementia (Ishii and Iadecola, 2015). These disorders are thought to affect the hypothalamic subnuclei differently, and typically only alter a subset of them (Bocchetta et al., 2015). Given the widespread documented homeostatic disturbances in ME/CFS, it is possible a number of these nuclei may be differentially affected in ME/CFS, though such relationships have yet to be investigated.

One nucleus of the hypothalamus that may be particularly implicated in ME/CFS pathophysiology is the paraventricular nucleus (PVN; Mackay and Tate, 2018). Located within the anterior-tuberal region, the PVN exerts control over most of the neuroendocrine axes and neuronal autonomic centres to regulate multiple homeostatic functions (Savić et al., 2022). Thus, changes to PVN circuitry may cause robust alterations to homeostasis (Rosenzweig et al., 2020) and disrupt functional integration between hypothalamic nuclei and processes relevant to ME/CFS pathophysiology. This includes interactions between the PVN and medial preoptic area, dorsomedial nucleus, lateral hypothalamus and suprachiasmatic nucleus to regulate the stress response (Gao and Sun, 2016), interactions with preoptic nuclei to initiate sleep (Jiang et al., 2021), and lateral and tuberomammillary nuclei to promote wakefulness (Ono et al., 2020). Disturbances to stress, sleep and wakefulness processes facilitated by the hypothalamus have been implicated in fatigue pathophysiology in other chronic conditions such as multiple sclerosis (Burfeind et al., 2016), but whether such relationships exist in ME/CFS have yet to be examined.

At present, there is limited consensus on the CNS disturbances present in adults and adolescents with ME/CFS as ascertained from neuroimaging studies. Consistency among findings has been hampered by differences in sample sizes, underlying disease heterogeneity, and methodology (Almutairi et al., 2020). For instance, many of the brain region-specific volumetric differences in ME/CFS have been derived from voxel-based statistics, which are biased towards group differences that are highly localised in space (Almutairi et al., 2020). In addition, one variable of relevance when examining brain-behaviour relationships in ME/CFS is the influence of illness duration. Recent imaging studies provide evidence to suggest CNS disturbances in ME/CFS become more pronounced over time, such that regional white and grey matter volumes decrease with increasing disease duration (e.g. Barnden et al., 2011; Shan et al., 2016), and functional changes become more pronounced with increasing fatigue duration in regions such as the brainstem (Barnden et al., 2015, 2011). Nacul et al. (2020) suggested increased disease duration may be associated with a prolonged state of low-grade and systemic inflammation, leading to greater CNS disturbance over time. However, limited studies have examined or accounted for the influence of illness duration on metrics derived from neuroimaging, thus the contribution of illness duration to CNS disturbances in ME/CFS pathophysiology is undetermined.

To date, no study has comprehensively investigated the involvement of the hypothalamus and its subregions in either adults or adolescents with ME/CFS. This is largely due to the application of whole-brain or low spatial resolution functional imaging techniques that preclude the study of small structures such as the hypothalamus, and challenges in manual measurements of hypothalamic regional volumes from MRI (Bocchetta et al., 2015; Schindler et al., 2019). Replicable, unbiased measurements of the hypothalamus and its subregions have recently been facilitated following the development of automated, freely available tools. One such segmentation tool by Billot et al. (2020) divides the hypothalamic nuclei into five subunits based on visible anatomical landmarks with specific nuclear subgroupings (Makris et al., 2013), including: (*i*) the anterior-superior hypothalamus; (*ii*) the anterior-inferior hypothalamus; (*iii*) the superior tuberal hypothalamus; (*iv*) the inferior tuberal hypothalamus; and (*v*) the posterior hypothalamus. Validation studies have identified regional changes within these hypothalamus subregions have meaningful links to functional disturbances in clinical populations such as frontotemporal dementia (Shapiro et al., 2022), multiple sclerosis (Genç et al., 2023) and obesity (Thomas et al., 2019). In addition, a growing body of research suggests structural changes related to pathology and neuroinflammation can be observed in prodromal and early stages of neurological disease (Oestreich and O’Sullivan, 2022), including subcortical gray matter volumes (Han and Ham, 2021), making it a viable tool for studying potential hypothalamic disturbances in adolescents recently diagnosed with ME/CFS.

In the present study, our primary aim was to examine whether lateralised regional and whole hypothalamus volumes in adolescents with ME/CFS differed compared with age-matched healthy controls. Second, we aimed to investigate whether lateralised regional and whole hypothalamus volumes were associated with severity of self-reported fatigue symptoms in both groups. Finally, we aimed to examine whether these volumes were associated with reported illness duration in the ME/CFS group.

## Methods and Materials

### Participants

48 participants (25 ME/CFS and 23 controls) were recruited for the study (as previously described in detail in (Josev et al., 2021, 2020). Inclusion criteria were as follows: (a) adolescents aged 13–18 years diagnosed with ME/CFS through a specialized tertiary hospital ME/CFS Clinic using the Canadian Consensus Criteria adapted for paediatrics (illness duration ≥3 months) (Jason et al., 2006), and; (b) healthy adolescent controls aged 13–18 years with no history of ME/CFS or other chronic illnesses.

### Procedure

The full procedure for this study has been described elsewhere (Josev et al., 2021, 2020). Briefly, participants completed a series of standardised questionnaires in the week prior to the assessment day at the hospital, administered via the online platform REDCap (version 5.10.2, Vanderbilt University, Tennessee, USA, 2014; Harris et al. 2009). On the assessment day, participants underwent a series of assessment measures including cognitive testing and an MRI scan. The measures relevant to the current study are described below.

### Fatigue measures

Fatigue was assessed using the *Pediatric Quality of Life Multidimensional Fatigue Scale* (PedsQL_TM_-MFS) Child Report (Varni et al., 2002), a well-validated measure of fatigue in paediatric patients with chronic illness (Crichton et al., 2015), including adolescents with ME/CFS (Knight et al., 2015). The 18-item PedsQL-MFS comprises 3 Subscales: (1) General Fatigue (6 items, e.g., ‘‘I feel tired’’; ‘‘I feel physically weak”), (2) Sleep/Rest Fatigue (6 items, e.g., ‘‘I feel tired when I wake up in the morning’’; ‘‘I rest a lot’’), and (3) Cognitive Fatigue (6 items, e.g., ‘‘It is hard for me to keep my attention on things’’; ‘‘It is hard for me to remember what people tell me’’). Items were reverse-scored and linearly transformed to a 0-100 scale (0 = 100, 1 = 75, 2 = 50, 3 = 25, 4 = 0), so that *higher* PedsQL-MFS scores indicate *fewer* symptoms of fatigue. Sum of all answered items from each scale provide a Total Fatigue Score, which was used for subsequent analysis.

### Illness duration

During consultation prior to the study visit, the treating paediatrician collected information about time since symptom onset on a 4-point scale of 1) 3-6 months; 2) 7-12 months; 3) 13-24 months, and; 4) >24 months, based on parent interview about illness characteristics including time since symptom onset to diagnosis. These four categories were used as an estimate of illness duration. For further detail about this measure, see Josev et al. (2021).

## Imaging

### Image acquisition

MRI data were acquired on a 3T Siemens Tim Trio research scanner (Siemens Medical Solutions, Erlangen, Germany) at the Royal Children’s Hospital, Melbourne, Australia. As part of the neuroimaging protocol, T1-weighted scans (3-dimensional motion-corrected multi-echo MPRAGE Magnetisation Prepared Rapid Gradient Echo; TR = 2530 ms, TE1 = 1.74 ms, TE2 = 3.60 ms, TE3 = 5.46 ms, TE4 = 7.32 ms, flip angle = 7.0 degrees, phase encoding direction = A >> P, coverage = whole brain including cerebellum, no. slices = 176, matrix = 208 × 246 × 176, resolution = 1 mm isotropic) were collected.

### Preprocessing

First, the neuroimaging data used in the study were organized using the Brain Imaging Data Structure (BIDS), version 2.1.6 (Gorgolewski et al., 2016). MRIQC version 22.0.6 (Esteban et al., 2017) was used to perform automated image quality control before analysis. Three output image quality metrics (ICMs) were used to evaluate the image quality; total signal-to-noise ratio (SNR), entropy focus criterion (EFC), and coefficient of joint variation (CJV). Following suggestions provided by Nárai et al. (2022), all participants satisfied quality control and no data were excluded.

Preprocessing was performed using the standard *recon-all* pipeline in the FreeSurfer image analysis suite (version 7.1.1; http://surfer.nmr.mgh.harvard.edu/). This pipeline performs intensity non-uniformity correction and normalization, skull stripping, and registration to the fsaverage template (based on the MNI305 template; Evans et al., 1992). Intracranial volume (ICV) was calculated for each participant using the estimated Total Intracranial Volume (eTIV) output from the *recon-all* pipeline (Buckner et al., 2004).

### Regional hypothalamus volume calculations

To extract the volumes of the hypothalamus and its five subregions (anterior-inferior, anterior–superior, inferior tubular, superior tubular and posterior), we used the (Billot et al., 2020) segmentation tool. Left and right hemisphere hypothalamus volumes were calculated by summing the five sub-regional volumes on the left and right, respectively. Subregions are depicted in *Figure 1*. Subnuclei within each segmentation are listed in *Supplementary Table 1*. Hypothalamic segmentations were visually examined by a single rater (HB) to ensure the hypothalamus and its subunits were correctly identified and segmented, and to exclude the presence of outliers. Visual inspection determined all segmentations to be suitable for analysis.

**Fig. 1.**
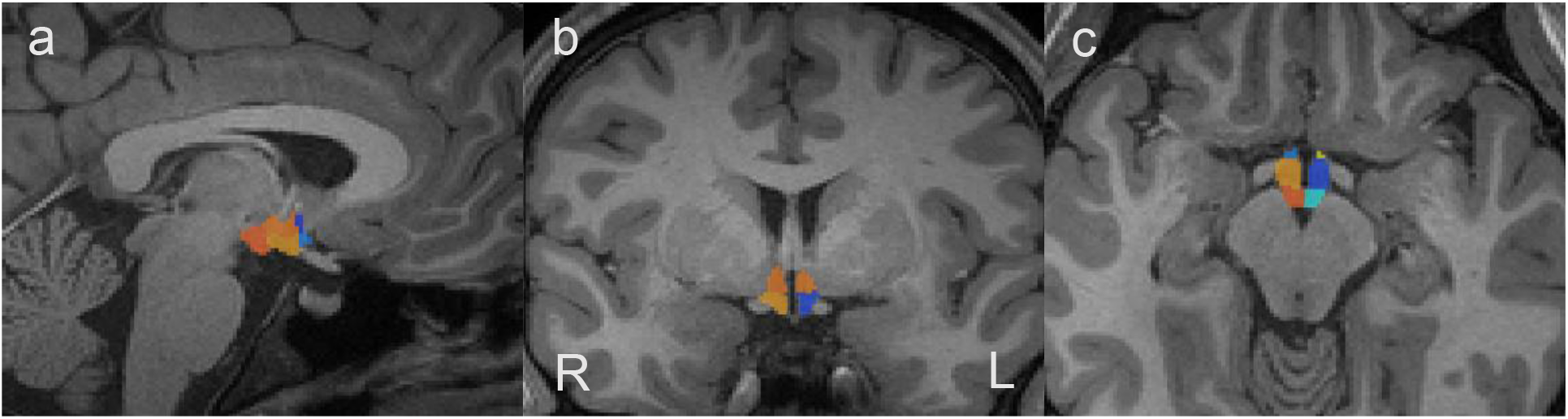
Sagittal (a), coronal (b) and axial (c) views of an example hypothalamus segmentation obtained from a participant. (a) right anterior-superior (dark blue), right anterior-inferior (light blue), right inferior tubular (yellow), right superior tubular (orange) and right posterior (dark orange); (b) superior tubular (light orange), right inferior tubular (dark yellow), left inferior tubular (blue); (c) right posterior (dark orange), right superior tubular (light orange), right anterior-inferior (light blue), left anterior-inferior (light yellow), left superior tubular (dark blue), left posterior (turquoise). Coronal and axial images are right-left reversed as per acquisition (R→L).

### Statistical analysis

All statistical analysis was performed using R Statistical Software (version 4.2.1; R Foundation for Statistical Computing, Vienna, Austria), run in RStudio (version 2022.07.2; http://www.rstudio.com/). Due to the modest sample size, a Bayesian analysis framework was adopted to provide accurate estimates of evidence of group differences independent of sample size (Makowski et al., 2019a). When possible, the descriptive features of the posterior probability distribution are reported: the Median of Posterior Distribution, which represent the point estimates for each parameter; Credible Intervals (CI), which represents uncertainty around these point estimates, as indicated by 95% highest density intervals; Probability of Direction (*pd*), where >97.5% *pd* was considered as the effect likely existing (corresponding to a two-sided *p*-value of .05), and; percentage in Region of Practical Equivalence (% in ROPE), where values below <2.5% were considered probably significant. Interpretation of % *pd* and % in ROPE were based on the reference values and descriptive features provided by Makowski et al. (2019a).

For all analyses, when possible, Bayes Factors (BF_10_) are reported, which are ratios providing the relative evidence of one “model” over another (Makowski et al., 2019a). In this case, these values correspond to relative strength of evidence for group differences and fatigue interactions in hypothalamic subregion volumes. BF_10_ values above 1 indicate increasing evidence in favour of the alternative hypothesis (i.e. that group differences or interactions between fatigue scores and regional hypothalamus volumes exist), with values between 1 and 3 said to provide ‘anecdotal evidence’ in favour of the alternative (i.e. weak evidence that may not necessarily be reliable) and values above 3 said to provide increasing evidence from moderate to strong in favour of the alternative hypothesis. BF_10_ values between 0.3 and 1.0 are typically interpreted as not offering conclusive evidence for or against the H_0_, and values below 0.3 indicate evidence in favour of the null hypothesis (Makowski et al., 2019a). Bayes analyses were performed using the *rstanarm* (Goodrich et al., 2020), *bayestestR* (Makowski et al., 2019b) and *brms* (Bürkner, 2021, 2018, 2017) packages in R. Given the functional asymmetry of hypothalamic nuclei (Kiss et al., 2020), statistical analyses were performed separately for lateralised total and regional hypothalamus volumes.

### Demographics and fatigue severity

Group differences in participant demographics (age and sex, according to patient medical record) and fatigue severity (i.e., PedsQL-MFS Total Fatigue scores) were evaluated using Bayesian Independent Samples T-tests (age and Total Fatigue scores) and a Bayesian test of association (sex). Note the sample characteristics of the groups have been previously examined using frequentist statistical methods (Josev et al., 2020), but were re-analysed here using Bayesian statistics for consistency within the current transcript.

### Corrected regional volumes comparison between ME/CFS and controls

To examine whether there were differences in regional hypothalamus volumes between the ME/CFS group and controls, we examined whether group status could predict regional hypothalamus volumes after controlling for age, sex and eTIV. Bayesian linear regression (estimated using MCMC sampling with 10 chains of 5000 iterations and a warmup of 1000) using the *rstanarm* package (Goodrich et al., 2020) were fitted to predict regional hypothalamus volumes with group, controlling for sex, age and eTIV (formula: *regional hypothalamus volumes ∼ group + sex + age + eTIV)*. Priors over parameters were set as normal distributions.

### Fatigue interactions with regional hypothalamus volumes

To examine whether regional hypothalamus volumes were associated with fatigue severity, we fitted Bayesian linear models (estimated using Hamiltonian Markov Chain Monte Carlo [MCMC; Carpenter et al. (2017)] sampling with 10 chains of 5000 iterations and a warmup of 1000) for the ME/CFS and control groups separately to examine whether whole and regional hypothalamus volumes could predict Total Fatigue scores when controlling for age, sex and eTIV (formula: *regional hypothalamus volumes ∼ Total Fatigue scores + sex + age + eTIV*) using the *rstanarm* package (Goodrich et al., 2020). Priors over parameters were set as normal distributions.

### Effects of illness duration on regional hypothalamus volumes in ME/CFS

In order to determine whether regional hypothalamus volumes were associated with illness duration, a Bayesian quantile (ordinal) regression model controlling for age, sex and eTIV was applied using the *brms* package (Bürkner, 2021, 2018, 2017). Parameters were estimated via MCMC with 10 chains each of 1000 warm-up and 10000 actual samples. Posterior predictive distributions were inspected to ensure adequate fit and the posterior predictive probability (*PPP*) computed for each model, with 0.5 < *PPP* < 0.95 considered to indicate adequate fit (Gelman et al., 2013).

## Results

### Demographic data

Descriptive characteristics for demographic variables and Total Fatigue scores are provided in *Table 1*. Illness duration data was missing for two ME/CFS participants (N = 23) and one ME/CFS participant did not complete the PedsQL-MFS due to fatigue (N = 24). There was no evidence of an age difference between groups (median [95% CI]: 0.09 [-0.66-0.92], BF_10_ = 0.295). In addition, the Bayesian Test of Association indicated there was no evidence for a difference in ratio of females to males between groups (BF_10_ = 0.61). However, there was very strong evidence to suggest the ME/CFS group scored differently from the control group on all PedsQL-MFS fatigue subscales (all BF_10_ values > 100). These findings were consistent with a previous analysis of this cohort using a frequentist approach (Josev et al., 2020).

**Table 1.** Participant characteristics and PedsQL-MFS subscale scores for the ME/CFS and control groups.

| Characteristic | ME/CFS | Controls | Median [95% CI] | $BF_{10}$ |
| --- | --- | --- | --- | --- |
| Age (years)<sup>1,3</sup> | 16.17 (13.42–18.92) | 16.33 (13.17–18.25) | 0.09 [-0.66–0.92] | 0.295 |
| Sex<sup>2,4</sup> |  |  |  | 0.61 |
| Female | 20 (80%) | 16 (70%) |  |  |
| Male | 5 (20%) | 7 (30%) |  |  |
| Total Fatigue Score<sup>1,3</sup> | 31 (0–64) | 75 (42–97) | -43.91 [-53.60–-33.61] | 9.66e+08 |
| Illness Duration<sup>2</sup> |  |  |  |  |
| 3-6 months | 5 (21.7%) |  |  |  |
| 7-12 months | 7 (30.4%) |  |  |  |
| 13-24 months | 6 (26.1%) |  |  |  |
| >24 months | 5 (21.7%) |  |  |  |

<sup>1</sup> Median (range); <sup>2</sup> N (%); <sup>3</sup> Bayesian Independent Samples T-test; <sup>4</sup> Bayesian Test of Association; CI = Credible Interval; $BF_{10}$ = Bayes Factor

### Effects of age, sex and eTIV on hypothalamus volumes

As age, sex and ICV are known to influence the size of hypothalamus volumes (Isiklar et al., 2022), a Bayesian Independent Samples T-test was used to assess the relationship between sex and volumetrics within each group, and Bayesian Pearson correlations were performed to look at the relationship between regional hypothalamus volumes and age, and eTIV. Bayesian Independent T-Tests revealed anecdotal evidence to suggest that, in the ME/CFS group, there was an effect of sex on whole left (median [95% CI] = 24.75 [-9.65, 66.30], BF_10_ = 1.17) and whole right (median [95% CI] = 35.13 [-3.41, 80.84], BF_10_ = 1.89) hypothalamus volumes. In the control group, there was anecdotal evidence for an effect of sex on whole right (median [95% CI] = 30.04 [-1.84, 66.70], BF_10_ = 2.10) volumes, but not the left (median [95% CI] = 10.08 [-22.31, 45.22], BF_10_ = 0.501). In both groups, Bayesian correlations did not identify a relationship between age and whole left (ME/CFS: median [95% CI] = -0.02 [-0.38, 0.33], BF_10_ = 0.435; controls: median [95% CI] = -0.19 [-0.52, 0.19], BF_10_ = 0.790) and right (ME/CFS: median [95% CI] = -0.11 [-0.45, 0.24], BF = 0.531; controls: median [95% CI] = -0.13 [-0.48, 0.23], BF_10_ = 0.569) hypothalamus volumes. However, there was moderate evidence of a positive correlation between eTIV and total left volumes in both groups (ME/CFS: median [95% CI] = 0.30 [-0.06, 0.58], BF_10_ = 6.47; controls: median [95% CI] = 0.41 [0.07, 0.68], BF_10_ = 5.13) and right volumes in both groups (ME/CFS: median [95% CI] = 0.46 [0.14, 0.70], BF_10_ = 12.92; controls: median [95% CI] = 0.41 [0.06, 0.68], BF_10_ = 6.53). Based on these findings, we included age, sex and eTIV in subsequent analyses to control for the potential impact of these variables on regional hypothalamus volumes.

### Differences in corrected regional hypothalamus volumes between ME/CFS and controls

Results from the Bayesian linear models (*Table 2*) provided very strong evidence (BF_10_ < 1/10) in favour of the null hypothesis, suggesting no differences in regional hypothalamus volumes between the ME/CFS and control group. Plots showing the data distribution of whole left and right hypothalamus volumes and marginal means with 95% confidence intervals are provided in *Figure 2*.

**Table 2.** Results from the Bayesian linear models examining the effect of group on regional hypothalamus volumes, controlling for age, sex and eTIV.

| Region | Hemisphere | Median [95% CI] | *pd* (%) | BF<sub>10</sub> |
| --- | --- | --- | --- | --- |
| Whole | Left | 0.28 [-23.77–24.41] | 51.36 | 0.06 |
|  | Right | -1.98 [-25.54–21.61] | 56.21 | 0.05 |
| Anterior-Inferior | Left | 0.58 [-2.05–3.25] | 67.38 | 0.07 |
|  | Right | -1.05 [-4.50–2.42] | 72.34 | 0.07 |
| Anterior-Superior | Left | -0.13 [-2.89–2.55] | 53.63 | 0.06 |
|  | Right | 0.42 [-2.99–3.82] | 59.40 | 0.06 |
| Posterior | Left | -1.98 [-9.98–5.06] | 71.59 | 0.07 |
|  | Right | -1.50 [-10.43–7.41] | 62.90 | 0.06 |
| Inferior Tubular | Left | 1.16 [-8.49–10.89] | 60.02 | 0.06 |
|  | Right | -0.33 [-9.18–8.42] | 52.51 | 0.05 |
| Superior Tubular | Left | 0.52 [-8.88–9.85] | 54.44 | 0.06 |
|  | Right | 0.56 [-7.43–8.53] | 55.66 | 0.06 |

Median = Median of Posterior Distribution; CI = Credible Intervals; *pd* = Probability of Direction; BF<sub>10</sub> = Bayes Factor

**Table 3.**
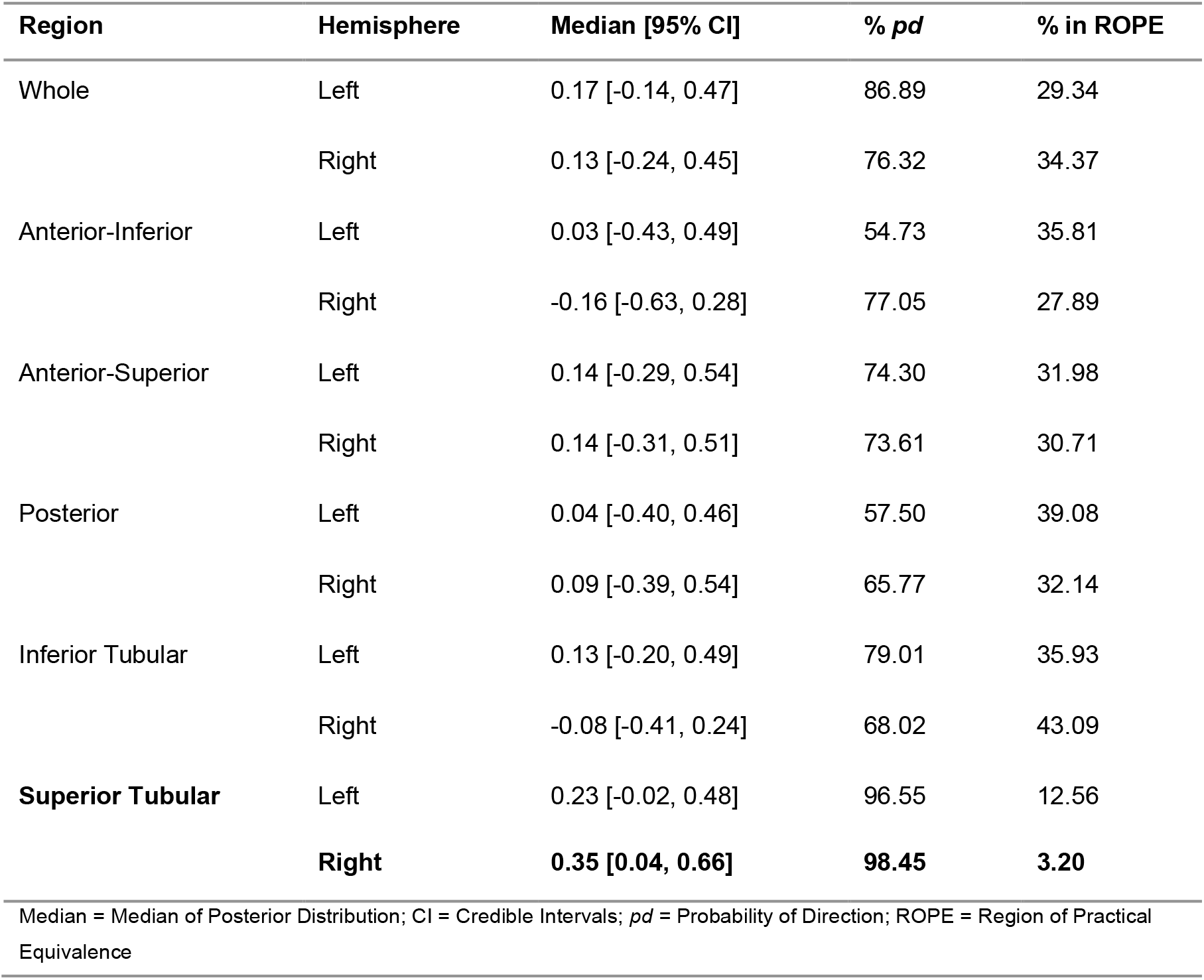
Results from Bayesian Regression Model examining whether illness duration predicts regional hypothalamus volumes when controlling for age, sex and eTIV.

| Region | Hemisphere | Median [95% CI] | % *pd* | % in ROPE |
| --- | --- | --- | --- | --- |
| Whole | Left | 0.17 [-0.14, 0.47] | 86.89 | 29.34 |
|  | Right | 0.13 [-0.24, 0.45] | 76.32 | 34.37 |
| Anterior-Inferior | Left | 0.03 [-0.43, 0.49] | 54.73 | 35.81 |
|  | Right | -0.16 [-0.63, 0.28] | 77.05 | 27.89 |
| Anterior-Superior | Left | 0.14 [-0.29, 0.54] | 74.30 | 31.98 |
|  | Right | 0.14 [-0.31, 0.51] | 73.61 | 30.71 |
| Posterior | Left | 0.04 [-0.40, 0.46] | 57.50 | 39.08 |
|  | Right | 0.09 [-0.39, 0.54] | 65.77 | 32.14 |
| Inferior Tubular | Left | 0.13 [-0.20, 0.49] | 79.01 | 35.93 |
|  | Right | -0.08 [-0.41, 0.24] | 68.02 | 43.09 |
| **Superior Tubular** | Left | 0.23 [-0.02, 0.48] | 96.55 | 12.56 |
|  | **Right** | **0.35 [0.04, 0.66]** | **98.45** | **3.20** |

Median = Median of Posterior Distribution; CI = Credible Intervals; *pd* = Probability of Direction; ROPE = Region of Practical Equivalence

**Fig. 2.**
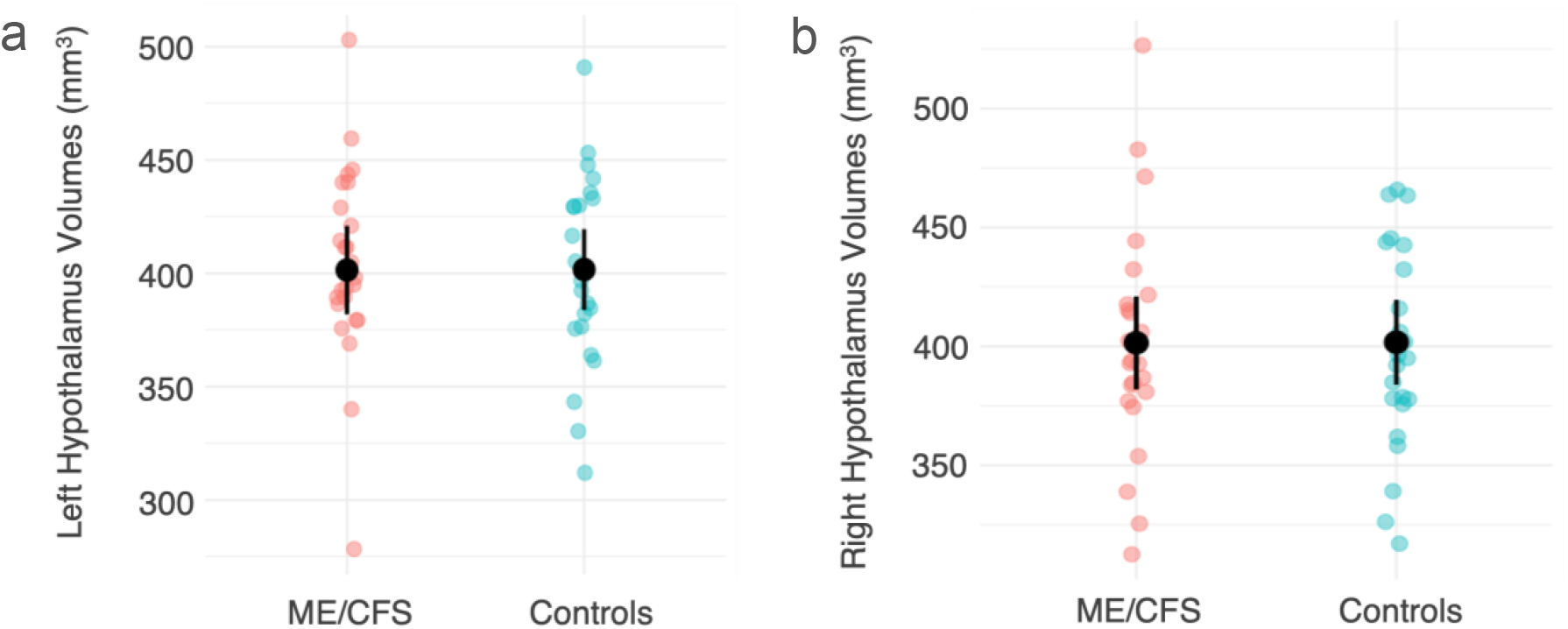
Jitter plots showing raw (uncorrected) whole left (a) and right (b) hypothalamus volumes for each group and their estimated marginal means with corresponding 95% upper and lower confidence intervals (bars).

### Association between regional hypothalamus volumes and fatigue in ME/CFS and controls

Results from the Bayesian linear models (*Figure 3*) provided anecdotal evidence for a negative linear relationship likely existing between Total Fatigue and right anterior-superior volumes in the ME/CFS group (median [95% CI]: -0.14 [-0.27–-0.02], *pd* (%) = 98.84, BF_10_ = 1.48), which was not present in the control group (median [95% CI]: 0.06 [-0.12–0.24], *pd* (%) = 75.59, BF_10_ = 0.11). In the ME/CFS group, this had undecided significance (21.32% in ROPE). Plots of this interaction is provided in *Figure 3*. No other relationships between lateralised regional or whole hypothalamus volumes and Total Fatigue scores were observed in either the ME/CFS or control groups.

**Fig. 3.**
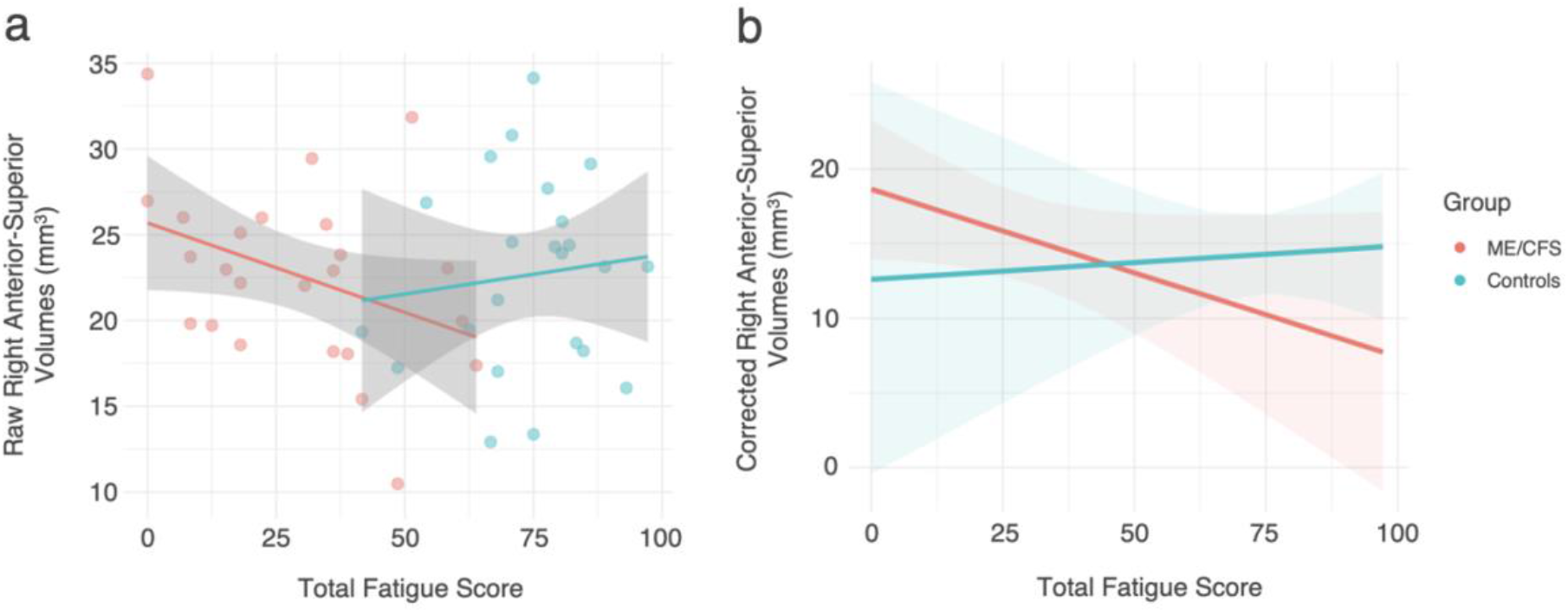
(a) Scatter plot showing relationship between raw (uncorrected) right anterior-superior volumes and Total Fatigue scores by group; (b) marginal means plot showing relationship between corrected right anterior-superior volumes and Total Fatigue by group. Note higher subscale scores indicate *fewer* symptoms of fatigue.

### Association between regional hypothalamus volumes and illness duration in ME/CFS

Results from the analysis testing for an association between regional hypothalamus volumes and the order of illness duration categories in the ME/CFS group provided strong evidence that illness duration did not predict regional hypothalamus volumes across the majority of subregions (median of posterior distribution range: -0.16–0.23, % *pd* range: 54.73–96.55, % in ROPE range: 12.56–43.09). However, the effect of illness duration on right superior tubular volumes had a 98.45% probability of being positive (median [95% CI] = 0.35 [0.04– 0.66]), but this had undecided significance (3.2% in ROPE). *PPP* sat just outside (−0.01) of the recommended threshold described by (Gelman et al., 2013). Plots showing the relationship between corrected and raw (uncorrected) right superior tubular volumes and illness duration are provided in *Figure 4*.

**Fig. 4.**
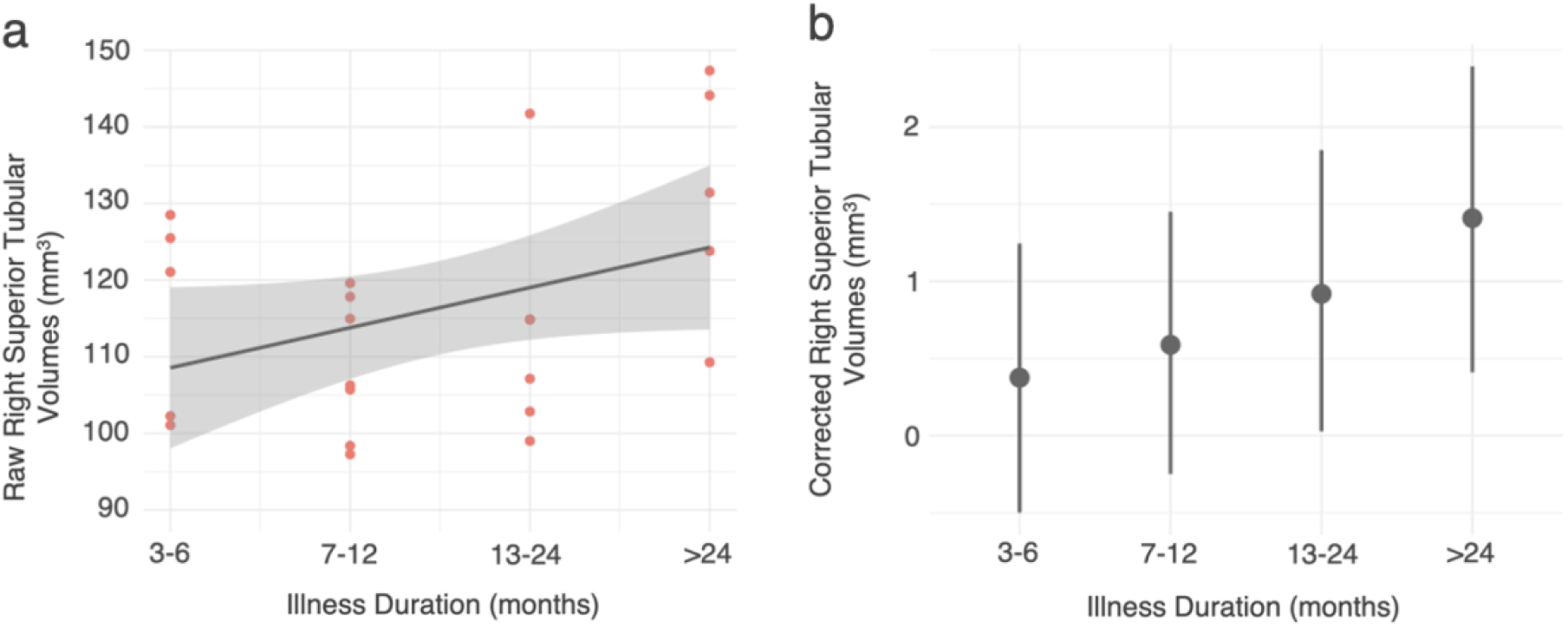
(a) Jitter plot showing raw (uncorrected) right superior tubular volumes for the ME/CFS group with increasing illness duration, and (b) plot showing right superior tubular volumes corrected for age, sex and eTIV over illness duration with 95% Credible Interval bars.

## Discussion

To our knowledge, this is the first study to explore whether volumetric changes within lateralised, functionally-distinct regions of the hypothalamus are present in adolescents with ME/CFS compared with matched healthy controls, and examine whether these volumes are associated with self-reported fatigue symptoms and illness duration.

### Hypothalamus volumes comparison

The primary observation from this study is that no differences in lateralised regional or whole hypothalamus volumes between the ME/CFS and matched control groups were identified. These findings suggest gross macrostructural changes to hypothalamic structure may not occur in adolescents with ME/CFS, despite the documented evidence of homeostatic disturbances in ME/CFS (Marks, 2023). However, as this study was conducted on a cohort of adolescents who were only recently diagnosed with ME/CFS, it is possible differences may emerge with increasing illness duration beyond two years. As such relationships could not be determined based on the clinical demographics in the current study, longer-term follow up studies would be beneficial in determining whether group differences in regional hypothalamus volumes emerge over time.

In addition, another consideration is patient diversity. ME/CFS is a heterogeneous condition where patients experience a wide variety of comorbidities and/or secondary symptoms at time-varying severities (Carruthers et al., 2011). This is particularly relevant to the present study, as recent research has identified ME/CFS patients can experience varying degrees of autonomic, neuroendocrine, and immunological disturbances (Murga et al., 2021), affecting the pattern and severity of homeostatic dysfunction. Consequently, looking at between-group differences may have precluded our ability to identify and compare subgroups or potential phenotypes of ME/CFS who may have differential, more pronounced, and/or fluctuating symptoms of homeostatic dysfunction. While standardised measures to assess homeostatic variability and severity in ME/CFS are limited, the need to document such individual differences in this population is gaining awareness (e.g. McDonald et al., 2022). Until such work is possible, replicable analyses such as those used in the current study provide utility in building a broader picture across cohorts, facilitating better understanding of the differing patterns of CNS disturbances relating to symptomatology in ME/CFS.

### Regional hypothalamus volumes and fatigue interactions

While no evidence for volumetric differences between groups was identified, there was some evidence of a weak negative linear relationship between right hemisphere anterior-superior volumes and greater fatigue symptoms in the ME/CFS group. This interaction was not present in the control group, suggesting the relationship between greater right anterior-superior volumes and greater fatigue severity was specific to ME/CFS.

The anterior-superior hypothalamus comprises the anterior portion of the nucleus of interest, the PVN. The interaction between the PVN, pituitary and adrenal glands comprising the hypothalamus-pituitary-adrenal (HPA) axis plays a fundamental role in regulating homeostasis via cortisol secretion, which serves to mobilise glucose reserves for energy and modulate inflammation in response to stress (Hannibal and Bishop, 2014). As low circulating cortisol in other clinical conditions is typically characterised by debilitating fatigue, impaired HPA axis functioning has long been considered in ME/CFS pathophysiology (Tanriverdi et al., 2007). Evidence of HPA axis disturbances in ME/CFS comes from multiple observations in both adult and adolescent ME/CFS patients of systemic hypocortisolism, enhanced corticosteroid-induced negative feedback, attenuated diurnal variation, and reduced responsivity to challenge (Tomas et al., 2013). However, investigations into HPA axis function have not always been consistent. Contradictory results have arisen from differences in diagnostic criteria, heterogenous patient populations, and methods used to test HPA axis function (Tanriverdi et al., 2007). Thus, the contribution of PVN disturbances to ME/CFS fatigue pathophysiology has yet to be determined. A follow-up study in a larger, longitudinal sample looking at PVN volumes, severity of HPA axis dysfunction and fatigue would be highly beneficial to understand whether such relationships are driving the findings observed in this study.

Increased subcortical volumes in the context of chronic illness or pathology are suggested to be related to neurogenic inflammation or glial cell genesis, cell size increases, changes in cortical synaptic connectivity or changes in blood flow (Chen et al., 2019; Trachtenberg et al., 2002). In adults with ME/CFS, a recent study identified significantly larger volumes in the brainstem compared to healthy controls (Thapaliya et al., 2023) which, taken with our study, suggest increased volumes in autonomic regions may be implicated in ME/CFS pathophysiology. Another recent study also identified increased gray matter volumes in posterior cingulate cortex, precuneus and superior parietal lobule positively correlated with self-reported fatigue in adults recently discharged from hospital following infection with SARS-CoV-2, but were still experiencing symptoms of fatigue (Hafiz et al., 2022). While hypothalamus volumes were not measured as part of this study, these results suggest limbic involvement in fatigue symptoms following acute SARS-CoV-2. As SARS-CoV-2 is becoming increasingly recognised as a trigger of ME/CFS (Salari et al., 2022) and post-acute COVID-19 syndrome demonstrates significant symptom overlap with ME/CFS (Marks, 2023), it is possible there is a pathophysiological substrate amongst these findings and the current study. However, further examination comparing ME/CFS, post-acute SARS-CoV-2 patients and healthy controls is required.

A further consideration is that, in addition to the PVN, the anterior-superior hypothalamus also comprises the preoptic area. This nucleus plays a crucial role in coordinating sleep and body temperature (Rothhaas and Chung, 2021), functions both typically affected in ME/CFS (McCarthy, 2022). Despite this, few have implicated the role of the preoptic area in ME/CFS symptomatology or general fatigue pathophysiology, except in animal models (e.g. Soares et al., 2007). While the potential involvement of the preoptic nucleus in the pathophysiology of fatigue in ME/CFS should not be discounted, there is currently greater evidence pertaining to the involvement of the PVN in ME/CFS, as measured by HPA axis and autonomic nervous system dysfunction. However, closer neuroanatomical investigations of the PVN and preoptic nucleus respectively would yield more conclusive results.

### Regional hypothalamus volumes and illness duration

In addition to fatigue interactions, based on the median values changing in a consistent way across the order of the illness duration categories, we found tentative evidence of a positive relationship between right superior tubular volumes and illness duration in the ME/CFS group.

The superior tubular contains the lateral portion of the PVN, in addition to the dorsomedial nucleus and the lateral hypothalamus, which play important roles in cognition and energy homeostasis (Arrigoni et al., 2019). The superior tubular region is densely connected with limbic structures such as the anterior cingulate cortex, amygdala, hippocampus and nucleus accumbens to regulate cognitive processing, sleep, wakefulness and arousal stability (Boes et al., 2018; Fujita et al., 2017). This network of limbic regions has been commonly implicated in the pathophysiology of fatigue (Kohl et al., 2009; Pellicano et al., 2010; Staud et al., 2018), and functional imaging studies have also identified neuroinflammation (Nakatomi et al., 2014) and reduced cerebral perfusion (Li et al., 2021) within the limbic system of adults with ME/CFS. This has led some researchers to suggest limbic system dysfunction originating from hypothalamic disturbances contribute to ME/CFS pathophysiology (Mackay and Tate, 2018). Such investigations have yet to be empirically tested and are beyond the scope of the current study, but warrant attention in future research.

### Additional observations

Interactions between Total Fatigue scores and illness duration were observed in the right anterior-superior and right superior tubular hypothalamus, respectively. These regions are directly adjacent to one another and contain the anterior and superior divisions of the PVN. The PVN was identified as the nucleus of interest for this study, and increased volumes within this region associated with fatigue and illness duration suggest there may be some association with PVN dysfunction in pathophysiology. However, as described prior, the anterior-superior and superior tubular regions also contains many reciprocal connections between limbic regions (Boes et al., 2018), and are crucially involved in regulating a number of other processes such as sleep and energy homeostasis (Fujita et al., 2017), so it is undetermined whether these interactions are linked to aberrant functioning within other systems. Therefore, closer investigation of HPA axis disturbances, PVN function and ME/CFS symptomatology is required to draw more substantiated conclusions.

Notably, interactions with fatigue severity and illness duration were each observed in the right hemisphere hypothalamus. While research in this space is limited, a recent pilot study by Manca et al. (2021) identified a trend towards increased functional connectivity between the right hemisphere hypothalamus and regions of the salience network following exertion in adults with ME/CFS, which was absent in the left hemisphere. Functional asymmetries in the hypothalamus have long been documented (Kiss et al., 2020), where the right hypothalamus is more dominant in regulating responses to stress (Xavier et al., 2009). There is also significant right-left asymmetry of neuroendocrine-gonadal function within the hypothalamus, particularly in the female sex (Kiss et al., 2020), that may be relevant to pathophysiology. Female sex is one of the most consistent predictive risk factors of a ME/CFS diagnosis (Lacerda et al., 2019), and females have an increased risk of ME/CFS onset during late adolescence compared to children under the age of 12 (Bakken et al., 2014). These factors point toward neuroendocrine involvement in ME/CFS pathophysiology (Thomas et al., 2022), and disturbances to hormones secreted via the hypothalamus-pituitary-gonadal axis have long been implicated in fatigue (Kaltsas et al., 2010) and ME/CFS pathophysiology (Martinkovich et al., 2014). Thus, further research examining the involvement of the right hemisphere hypothalamus in ME/CFS neuroendocrine disturbances would be beneficial.

## Limitations

There are limitations to consider with this study. First, this study contained no assessment of the severity or pattern of homeostatic disturbances in either the ME/CFS or control groups, limiting our ability to directly examine the contribution of homeostatic dysfunction to regional hypothalamus volumes. Second, another notable limitation is the modest sample size. While a Bayesian framework can provide a conservative estimate for the relative evidence for an alternative hypothesis over the null even in small sample sizes (Jarosz and Wiley, 2014), this still likely contributed to the majority of our findings only providing anecdotal evidence in favour of the alternative hypothesis. Thus, our results must be interpreted as preliminary. This is particularly relevant for the illness duration variable where each time category only represented a few participants, making our results more susceptible to the effects of outliers.

A further limiting factor is the categorical nature of the illness duration variable, which means important descriptive information about their specific illness duration is missing – for example, category three covers 13-24 months illness duration, and each participant could be sitting at any duration within the given timeframe. Statistically, we employed the most appropriate technique for this data, which was to estimate the likelihood that the value of the outcome changed over binaries in a specific order. We acknowledge the limitations our data and this analysis, but report the preliminary results here as useful direction for future investigation. Finally, while the segmentation tool by Billot et al. (2020) provides a useful and replicable tool for studying specific nuclei subgroupings within the hypothalamus in ME/CFS, we were not able to examine relationships between specific nuclei and clinical measures (i.e. fatigue symptoms and illness duration). This is particularly relevant to our understanding of the contribution of PVN disturbances to the interaction results, where further region of interest studies are needed. Until more specific neuroanatomical studies can be conducted, this study provides a useful stepping point for future examination of hypothalamus disturbances in adolescents with ME/CFS.

## Conclusions

To our knowledge, this is the first study to examine differences and clinical relationships in regional hypothalamus volumes in either the adult or adolescent ME/CFS population. Our results suggest regional hypothalamus volumes may not be affected in ME/CFS during adolescence, but further research is needed to establish whether our ability to observe group differences was affected by the severity or pattern of homeostatic disturbances in the ME/CFS group. We did, however, observe a weak negative linear relationship between right anterior-superior volumes and Total Fatigue scores in the ME/CFS group that was absent in the control group, suggesting a potential association between greater fatigue symptoms and larger right anterior-superior volumes in ME/CFS. Finally, we also identified preliminary evidence to suggest superior tubular volumes may be larger after longer periods of illness duration (i.e. >24 months compared to less than 6), warranting further exploration in a larger, longitudinal sample. Understanding the contribution of hypothalamic disturbances to ME/CFS may aid in the development of evidence-based treatment options for the condition, and this study has implications for the need to document symptom heterogeneity and fluctuation in ME/CFS. To this end, replicating this analysis in a larger, longitudinal sample of both adults and adolescents with ME/CFS of varying illness duration with more detailed exploration of homeostatic disturbances would be highly beneficial, which can be facilitated by our automated, replicable approach.

## Data Availability

All data produced in the present study are available upon reasonable request to the authors.

https://github.com/DevelopmentalImagingMCRI/mecfs

## Acknowledgements

This study was funded by ME Research UK (SCIO charity number SCO36942, http://www.meresearch.org.uk/) and Judith Jane Mason & Harold Stannett Williams Memorial Foundation (Mason Foundation, ABN96140017725). Hollie Byrne is supported by the Melbourne Research Scholarship. This research was conducted within the Developmental Imaging and Neurodisability and Rehabilitation research groups at the Murdoch Children’s Research Institute, and the Children’s MRI Centre at the Royal Children’s Hospital, Melbourne, Victoria. This study was supported by the Murdoch Children’s Research Institute, the Royal Children’s Hospital, the Department of Paediatrics at The University of Melbourne and the Victorian Government’s Operational Infrastructure Support Program. We would like thank the CFS Rehabilitation Clinic and the Medical Imaging staff at the Royal Children’s Hospital for their assistance with this study, and the research participants and their families for generously donating their time to this research.

## Statements and Declarations

### Funding

This study was funded by ME Research UK (SCIO charity number SCO36942) and Judith Jane Mason & Harold Stannett Williams Memorial Foundation (Mason Foundation, ABN96140017725). Hollie Byrne is supported by the Melbourne Research Scholarship.

### Competing Interests

The authors have no relevant financial or non-financial interests to disclose. The funders had no role in the design of the study, in collection, analyses, or interpretation of data, in the writing of the manuscript, or in the decision to publish the results.

### Ethics approval

All procedures were conducted in accordance with the ethical standards of the institutional and/or national research committee and with the 1964 Helsinki declaration and its later amendments or comparable ethical standards. The study was approved by the Royal Children’s Hospital Human Research Ethics Committee (HREC 32233, 37200).

### Author Contributions

Conceptualisation: HB, EKJ, SJK, MLS; Formal analysis: HB, MLS; Data Curation: HB, EKJ; Writing – original draft preparation: HB; Writing – reviewing and editing: all authors; Supervision: EJK, SJK, MLS; Funding acquisition: EKJ, SJK, AS.

### Data Availability

The datasets generated during and/or analysed during the current study are not publicly available due to patient privacy and possible re-identification, but are available from the corresponding author on reasonable request. For replication in other datasets, the code used to run the statistical analysis has been uploaded online and can be viewed here: https://github.com/DevelopmentalImagingMCRI/mecfs

### Consent to participate

Informed consent was obtained from all individual participants and their parents included in the study.

---

**Supplementary Table 1.**
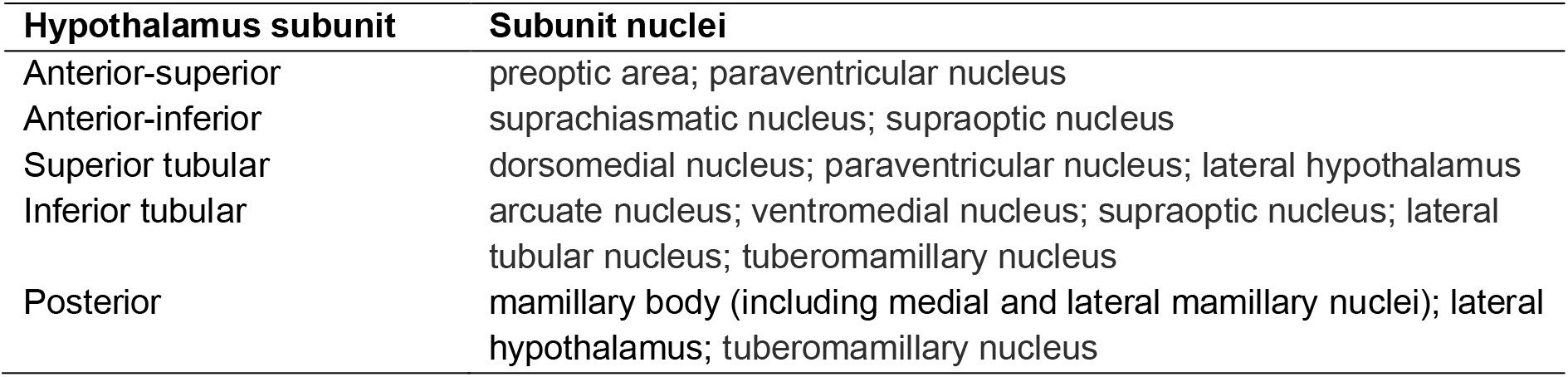
Table adapted from Billot et al. (2020). Grouping of hypothalamic subunit nuclei, based on Bocchetta et al. (2015) and Makris et al. (2013).

## Notes

### Competing Interest Statement

The authors have declared no competing interest.

### Author Declarations

The Human Research Ethics Committee (HREC 32233, 37200) of the Royal Children's Hospital gave ethical approval for this work.

