## Supplemental Table 1 for "Hypothalamus volumes in adolescent Myalgic Encephalomyelitis/Chronic Fatigue Syndrome: Impact of self-reported fatigue and illness duration"

**Supplementary Table 1.** Table adapted from Billot et al. (2020). Grouping of hypothalamic subunit nuclei, based on Bocchetta et al. (2015) and Makris et al. (2013).

| Hypothalamus subunit | Subunit nuclei |
|---|---|
| Anterior-superior | preoptic area; paraventricular nucleus |
| Anterior-inferior | suprachiasmatic nucleus; supraoptic nucleus |
| Superior tubular | dorsomedial nucleus; paraventricular nucleus; lateral hypothalamus |
| Inferior tubular | arcuate nucleus; ventromedial nucleus; supraoptic nucleus; lateral tubular nucleus; tuberomamillary nucleus |
| Posterior | mamillary body (including medial and lateral mamillary nuclei); lateral hypothalamus; tuberomamillary nucleus |
